# Reconciling neurocognitive and behavioral impulsivities through ecological assessment and multivariate modelling of cognitive control dynamics

**DOI:** 10.64898/2026.04.27.26351677

**Authors:** Andrea Imparato, Natacha Reich, Gregoire Riviere, Stephan Eliez, Christopher Graser, Maude Schneider, Corrado Sandini

## Abstract

Impulsivity is a core dimension of ADHD and a transdiagnostic risk factor for adverse outcomes. However, laboratory-based neurocognitive measures show weak convergence and limited prediction of real-world behavior. We tested whether repeated ecological assessment combined with response-time (RT) modeling improves impulsivity assessment. Sixty participants completed 1,347 smartphone-based Balloon Analogue Risk Task (D-BART) assessments alongside the Conners CPT-3. Linear mixed-effects models characterized RT as a function of objective risk and subjective uncertainty, while partial least squares identified shared D-BART/CPT-3 response patterns. RT slowed with increasing risk and uncertainty, distinguished clinical from control participants better than conventional metrics. D-BART latent scores predicted real-world hyperactivity/impulsivity with superior accuracy compered to CPT-3 scores. D-BART predictive accuracy further increased with repeated ecological sampling. These findings demonstrate that repeated ecological assessment combined with parsimonious RT modelling improves the construct validity, cross-paradigm convergence, and clinical relevance of neurocognitive measures of impulsivity beyond the current clinical gold standard.

## Introduction

Impulsivity is broadly defined as a predisposition toward rapid, unplanned actions that are misaligned with situational demands and executed without adequate consideration of their consequences [1]. It is a core dimension of Attention-Deficit/Hyperactivity Disorder (ADHD)[2], a highly prevalent neurodevelopmental disorder affecting approximately 5% of children [3, 4] and 2.5% of adults[4]. Beyond ADHD, impulsivity is commonly observed across a range of conditions, including disruptive behavior disorders, borderline personality disorder, bipolar disorder, and substance use disorders [1, 2, 5-9]. Emerging evidence suggests that impulsivity may not merely be a shared trait across disparate diagnoses but rather a transdiagnostic endophenotype contributing to their co-occurrence within individuals and families [10-13], a view supported by genetic studies demonstrating shared polygenic vulnerability across disorders characterized by poor impulse control [14-16]. Clinically, impulsivity is a strong longitudinal predictor of adverse outcomes, including substance misuse, obesity, academic failure, antisocial behavior, criminality, risky driving, self-injury, and impaired social and occupational functioning[12, 17-19] [20, 21]. Many of these adverse outcomes can be substantially mitigated through early identification and effective treatment[22], underscoring the need for robust tools to assess and monitor impulsivity across development and clinical contexts[23, 24].

Current pathophysiological models attribute impulsive behavior to insufficient top-down control from prefrontal regions over subcortical circuits supporting goal-directed behavior [13, 25]. This dysfunction can be assessed using neurocognitive paradigms probing inhibitory control [25]. The Conners Continuous Performance Test (CPT-3) [26] is widely used to assess attentional control, with impulsivity operationalized as failures to suppress reflexive responses, resulting in poor discrimination between target and non-target stimuli [27]. Related paradigms, including the Go/No-Go and Stop Signal Tasks, similarly assess suppression of prepotent responses [28, 29]. Higher-order aspects of impulsivity, including risk-reward evaluation and delay discounting, are assessed using paradigms such as the Iowa Gambling Task [30] and the Balloon Analogue Risk Task (BART)[31]. The BART provides a particularly useful dynamic index of impulsivity, requiring individuals to balance the potential gains of further balloon inflation against the risk of losing accrued rewards, when excessive inflation results in balloon explosion.

Despite their richness, current neurocognitive paradigms face two major shortcomings. First, behavioral measures of impulsivity often show weak convergence across tasks, raising the question of whether impulsivity reflects a unitary neurobiological construct or multiple distinct processes [25, 32]. Second, laboratory-based neurocognitive measures correlate only modestly with real-world impulsive behavior [33-38], strongly limiting their clinical utility [35]. We propose that these shortcomings may arise, at least in part, from two methodological features of conventional neurocognitive assessment: the reliance on single assessments performed under standardized laboratory conditions and the use of aggregate task outcomes that may provide limited insight into the cognitive processes generating behavior. We address each of these features in turn and evaluate whether doing so improves convergence across paradigms and prediction of clinically meaningful behavior.

The first methodological feature concerns how neurocognitive function is sampled [36, 37]. Impulse control has traditionally been conceptualized as a stable trait measured during a single standardized assessment [39]. However, growing evidence indicates that it fluctuates substantially as a function sleep[40], arousal [41-43], and other transient internal states [44], while real-world impulsive behavior is strongly shaped by environmental context [45]. Thus, real-world manifestations of impulsivity may depend less on peak performance under ideal testing conditions and more on the consistency of self-regulation across varying internal states and environments. Smartphone-based adaptations of paradigms such as the BART [33, 46]now enable repeated ecological assessment across everyday contexts [47]. Although previous studies have largely validated mobile versions against their laboratory counterparts [29, 35], it remains unknown whether repeated ecological assessment improves prediction of clinically meaningful behavioral outcomes beyond established laboratory measures such as the CPT-3. To address this question, we administered a smartphone-based BART to adolescents with ADHD, individuals with 22q11.2 deletion syndrome—a genetic population at elevated risk for ADHD [48] and healthy controls, all of whom completed gold-standard clinical and neurocognitive assessments.

The second methodological feature concerns how neurocognitive performance is quantified. Aggregate measures such as commission errors or balloon explosions provide limited insight into the cognitive mechanisms generating behavior, whereas response-time (RT) dynamics may provide a window into adaptive cognitive-control processes and potentially bridge performance across paradigms [49, 50]. For example, although the Stop Signal Task is typically conceptualized as a lower-order motor impulsivity task, dynamic RT analyses reveal adaptive slowing as stop-signal probability increases, reflecting strategic modulation of speed–accuracy trade-offs and engagement of top-down control [49, 50]. These findings align with dual-process theories of higher-order decision-making, which propose that low-risk decisions rely primarily on fast, automatic processes, whereas increasing risk and uncertainty recruit slower, deliberative control [51-54], an idea popularized as “*Thinking, Fast and Slow* [54]”. In the BART, this framework predicts that impulsive individuals should not only take greater risks but also exhibit reduced adaptive RT slowing as explosion probability increases, suggesting diminished engagement of deliberative control[52, 55-57]. Although increasingly informative, existing RT models remain computationally complex, and it remains unclear whether they can be translated into clinically useful individual-level measures that bridge neurocognitive paradigms or better predict real-world behavioral outcomes[52, 55-57]. To address this question, we developed a parsimonious linear mixed-effects framework for modelling BART RT dynamics that minimizes theoretical assumptions while leveraging the enhanced ecological validity afforded by repeated smartphone-based assessments.

We hypothesized that adaptive engagement of cognitive control would be reflected by RT slowing as objective explosion risk increased and as participants approached the decision to cash out, reflecting increasing subjective uncertainty. We further hypothesized that this dynamic shift in cognitive control would vary across both individuals and assessments, explain variability in traditional BART performance metrics, and capture a shared neurocognitive construct bridging BART and CPT paradigms, which we tested using multivariate partial least squares analysis. Finally, we tested whether combining repeated ecological assessment with dynamic RT modelling improved prediction of real-world behavioral impulsivity compared with the CPT-3, and whether any improvement reflected repeated ecological assessment rather than intrinsic differences between the two paradigms.

## Results

### Modelling Response Styles Through Response Time Dynamics in Clinical and Healthy Populations

A smartphone-based version of the BART was administered as part of a broader digital neurocognitive battery (including digital Trail Making and Corsi working memory tasks) in 59 participants. The sample comprised 40 individuals with recently diagnosed idiopathic ADHD, 10 participants with 22q11.2 deletion syndrome (22q11DS) recruited through the Swiss longitudinal cohort, and 9 healthy controls (HC) recruited among unaffected siblings of 22q11DS participants. Participants received smartphone notifications three times per day prompting completion of the neurocognitive battery. The 22q11DS and HC groups completed assessments over a 2-day period, whereas the ADHD group completed testing over a 30-day ecological protocol designed to capture real-world fluctuations in cognition, behavior, and sleep. This yielded a total of 1347 completed BART sessions, while number of per-participant sessions varied according to study duration: 2.67 ± 0.71 (HC), 2.80 ± 0.92 (22q11DS), and 32.38 ± 25.20 (ADHD). Each BART session consisted of 10 balloons. Participants earned one point per inflation while risking balloon explosion and loss of accumulated points. They could choose to “cash out” at any time. Explosion thresholds varied across trials (mean = 10 pumps, SD = 5), implying that balloon explosion risk increased with balloon inflation (BI), with BI of 10, representing a tipping point after which explosion became the most likely outcome of further inflation.

We first examined whether reaction time (RT) between successive decisions varied as a function of balloon inflation (BI), indexing objective explosion risk, while accounting for random variation across balloons (See methods for details). RT modeling was performed exclusively on successful trials culminating in point collection, allowing us to control for reverse causal associations between task performance and dynamic response patterns, and ensuring that directly comparable data were used to model both objective risk and subjective uncertainty. RT increased significantly as a function of BI (t = 24.128, p < .0001), indicating that participants progressively slowed their responses as objective risk increased (Figure-1-Panel-A1). In accordance with the Dual-Process Framework[51-54], this pattern suggests greater engagement of slower, deliberative processes under higher-risk conditions.

**Figure 1:**
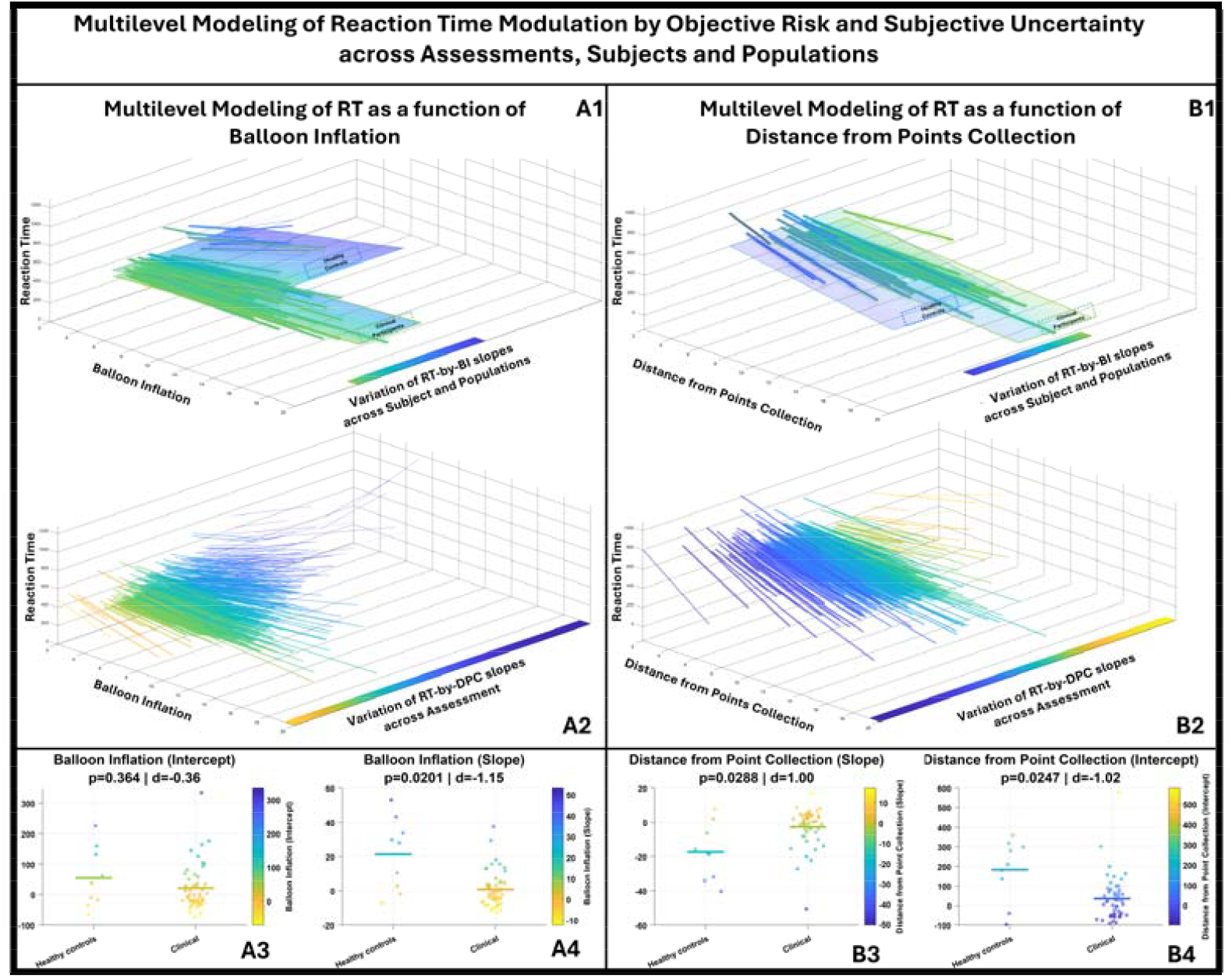
Multilevel modelling of Reaction-Time modulation by objective risk and subjective uncertainty across assessments, subjects and populations. **Panel A1/A2:** Multilevel modelling of Reaction-Time, (represented on the vertical Y axis) by Balloon-Inflation (represented on horizontal X axis), capturing variation in response style adaptation to objective risk (RT-by-BI slope), across assessments, subjects, and populations derived from mixed-effects modelling and represented along the Z axis. Across both Panels-A1 and A2, blue to yellow color coding, and positioning along the Z axis, encodes RT-by-BI slope values using a common scale, allowing direct comparison of trajectory steepness across assessment, subject, and population levels. **Panel-A1** represents variation in RT-by-BI slope across subjects and populations. The large surfaces represent the mean fitted trajectory for Healthy Controls (represented with dashed outlines) and Clinical Participants (represented with full outlines). Their position along the slope dimension and their color both reflect the mean population slope. Their width along the slope dimension corresponds to mean ± 2 standard deviations of the population slope distribution, thereby providing a visualization of between-subject variability within each population. The smaller ribbons superimposed on these population-level surfaces represent subject-level mean fitted trajectories. Each subject ribbon is positioned along the Z-slope axis and colored according to the participant’s mean slope across assessments, thus summarizing that participant’s average response style. The outline of subject-specific ribbon is colored according to the mean slope his respective population, with dashed and full outlines identifying Healthy Controls from Clinical Participants respectively, thus providing a representation of the extent to which individual participants differed from their respective population-based average. Within each subject-specific ribbon, semi-transparent internal lines represent that participant’s assessment-level fitted trajectories. Lines closer to the subject mean are less transparent, whereas lines farther from the subject mean are more transparent, allowing visualization of how repeated assessments are distributed around the participant’s average fit. **Panel-A2** captures assessment specific RT-by-BI slope variation. **Panel-A3:** Distribution of participant-level Reaction-Time by Balloon-Inflation intercept values, capturing RT under low-risk low-inflation conditions, compared across Healthy Controls and Clinical Participants. **Panel-A4:** Distribution of participant-level Reaction-Time by Balloon-Inflation slope values, capturing modulation of RT as a function of objective risk, compared across Healthy Controls and Clinical Participants. **Panel B1/B2:** Multilevel modelling of Reaction-Time, (represented on the vertical Y axis) by Distance from Point Collection (DPC represented on horizontal X axis), capturing variation in response style adaptation to subjective uncertainty (RT-by-DPC slope), across assessments, subjects, and populations derived from mixed-effects modelling and represented along the Z axis. The structure of Panels B1/B2 recapitulates that of Panels A1/A2 except that fitted trajectories are derived from the RT-by-DPC mixed-effects model and the x-axis represents Distance from Points Collection. **Panels B3–B4:** Same as panels A3–A4, but for random intercept (B3) and random slope (B4) estimates derived from the RT-by-DPC mixed-effects model.

Informed by recent findings[53], we next evaluated whether subjective confidence/uncertainty with regards to the optimal decisional outcome (inflate or collect) for a given level of objective risk, would also impact RT. We hypothesized that subjective uncertainty would peak immediately prior to the decision to collect points, that can be conceptualized as an uncertainty tipping point, at which the subjective appraisal of explosion risk outweighs the value attributed to further successful inflations. Subjective uncertainty was therefore operationalized using distance from point collection (DPC), reflecting the number of pumps remaining before cash-out. Consistent with predictions, RT was maximal immediately prior to point collection and decreased as DPC increased (t = –19.495, p < .0001). This finding supports differential engagement of fast-automatic versus slow-reflexive processes as a function of decisional uncertainty[53].

Next, we investigated whether modulation of RT by risk and uncertainty varied across individuals, by extending these RT models to include participant-level random intercepts and slopes (See methods for details). Likelihood ratio tests comparing models with and without participant-level random effects demonstrated significant improvements in model fit for both BI (χ^2^(3) = 2843.7, p < .0001), and DPC (χ^2^(3) = 3118.7, p < .0001), indicating substantial inter-individual variability in the extent to which objective risk and subjective uncertainty modulated RT. This variability reflects meaningful differences in dynamic cognitive-control engagement across participants. Finally, we examined whether modeling temporal fluctuations occurring across each individual’s repeated assessments would further improve model fit by extending the model to include session-level random intercepts and slopes. Inclusion of session-level random effects yielded additional significant improvements in model fit for both BI (χ^2^(3) = 3178.9, p < .0001) and DPC (χ^2^(3) = 2759.7, p < .0001), indicating that variability across game sessions independently contributed to RT dynamics.

Together, these findings demonstrate that RT modulation by risk and uncertainty is robust at the group level, varies substantially between individuals, and also fluctuates within individuals across repeated ecological assessments, supporting the presence of both trait-like and state-dependent components of impulse-control engagement (Figure-1-Panel-A2-B2).

### Comparing Response-Time Dynamics and Traditional BART Performance Metrics Across Samples

We next examined whether heterogeneity in response styles was linked to population-specific factors differentiating healthy controls (HC) from clinical participants (CP; ADHD and 22q11DS combined). To this end, population (HC vs CP) was explicitly modeled within a mixed-effects framework as a fixed effect and testing its interaction with balloon inflation (BI) and distance from point collection (DPC) (See methods for details). Results revealed a highly significant population-by-BI interaction (t = –4.6366, p < .0001), indicating a steeper increase in RT with increasing objective risk in HCs relative to CPs (Figure-1-Panel-A1). Similarly, a significant population-by-DPC interaction (t = 3.7899, p < .0002) demonstrated a stronger influence of subjective uncertainty on RT in HCs compared to CPs (Figure-1-Panel-B1). Within the Dual-Systems framework, these findings suggest that HCs show a more pronounced shift from fast-automatic to slow-deliberative processing as risk and uncertainty increase, whereas this adaptive cognitive-control shift appears attenuated in clinical participants.

We then investigated whether mixed-effects modeling could yield subject-specific indices of this cognitive-control shift. A model including balloon-level random intercepts and game-level random intercepts and slopes for both BI and DPC generated four dynamic response-style parameters: RT-by-BI intercept, RT-by-BI slope, RT-by-DPC intercept, and RT-by-DPC slope. We obtained participant-level estimates for each of those parameters by averaging assessment-specific parameters across sessions. For comparison, we also computed participant-level averages for commonly used traditional BART metrics: mean collected points (overall performance), proportion of exploded balloons, mean inflation at point collection (risk-taking propensity), mean RT, and RT standard deviation. Two-sample t-tests showed that HCs and CPs did not differ significantly on any traditional BART outcome measures, including mean points earned, mean collection point, or proportion of exploded balloons (see supplementary materials). Instead, HCs exhibited significantly higher RT-by-BI slope estimates (*d* = 1.15, p = .021, Figure-1-Panel-A4), reflecting greater slowing of RT as objective risk increased. This reduced modulation in CPs contributed to a smaller overall increase in mean RT relative to HCs (*d* = .93, p = .044). No significant group differences were observed in RT-by-BI intercepts (*d* = .36, p = .365), indicating comparable RTs under low-risk conditions. Regarding subjective uncertainty, HCs displayed significantly higher RT-by-DPC intercepts (*d* = 1.02, p = .024, Figure-1-Panel-B3), corresponding to longer RTs immediately prior to cash-out, and significantly lower RT-by-DPC slopes (*d* = 1.00, p = .028, Figure-3-Panel-B4), indicating sharper acceleration of RT as uncertainty decreased.

Together, these findings indicate that dynamic RT-based measures of cognitive-control engagement provide greater sensitivity in distinguishing clinical participants from healthy controls than conventional aggregate BART performance metrics.

### External Validation of D-BART against CPT-3 Neurocognitive Battery

After comparing the isolated diagnostic performance of RT and performance-based variables, we then investigated the added value of combining them together. Indeed, within the Dual-Systems Framework, insufficient cognitive control is defined both by an inability to flexibly shift between fast/automatic and slow/deliberative response styles according to situational demands, *and* by the resulting inefficient decisional outcomes. To more accurately capture such dysfunction, we integrated both traditional BART performance indices and dynamic RT-based metrics through multivariate Partial Least Squares (PLS) analysis, correlating them with nine age-and-gender-normed behavioral readouts derived from the gold-standard CPT-3 neurocognitive battery (Detectability, Omissions, Commissions, Perseverations, Hit Reaction Time (Hit RT), Hit RT Standard Deviation, Variability, Hit RT Block Change, and Hit RT Inter-Stimulus Interval (ISI) Change). PLS is specifically designed to identify latent components capturing shared covariance between two distinct variable sets and thus served two complementary purposes: (i) to characterize how dynamic and traditional BART metrics covary within the same instrument, and (ii) to determine whether their combined expression yields a latent signature of insufficient cognitive control that bridges across assessment paradigms, thereby providing evidence of external validity.

PLS analysis identified a first latent component (LC1) capturing a significant multivariate correlation between CPT-3 and D-BART response patterns (r = 0.37, p = 0.0037, Figure-2-Panel-D), accounting for 82% of shared covariance (Figure-2-Panel-A). Each latent component was associated with paired loading patterns—one for CPT-3 variables and one for D-BART variables—representing maximally correlated multivariate expressions across subjects. Reliability of individual variable contributions was assessed using bootstrap resampling (500 iterations). On the CPT-3 side, LC1 reflected globally reduced performance, with strong positive loadings for commission errors and impaired Detectability (*d*′), alongside fast reaction times (negative loading on Hit RT) and increased RT variability (Figure-2-Panel-B). This pattern is consistent with an impulsive response style characterized by poor discrimination and inefficient speed–accuracy regulation[58]. When interpreted through the Dual-Systems Model lens[49, 51, 53, 54], these findings may suggest an inability to differentially engage fast-automatic versus slow-reflexive systems, leading to inefficient modulation of speed–accuracy trade-offs and providing a signature of deficient cognitive control that may potentially bridge across different behavioral paradigms.

**Figure 2.**
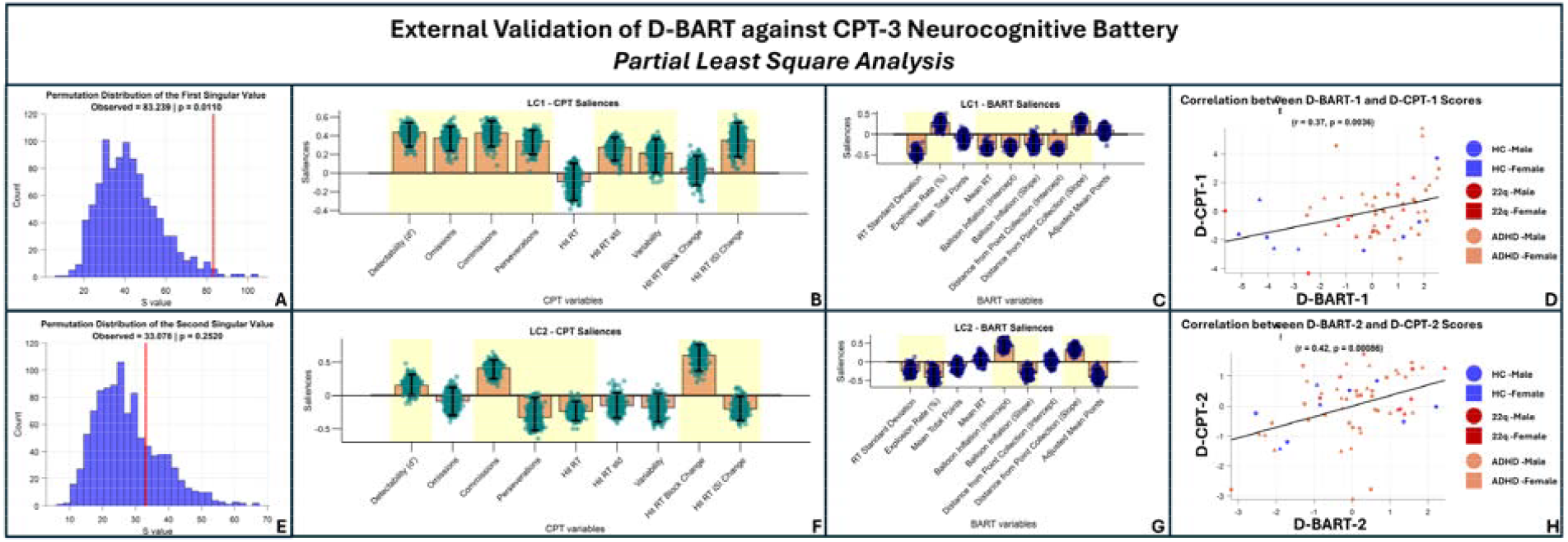
Partial Least Squares (PLS) analysis linking BART task and CPT-3 neurocognitive measures. **Panels A and E:** Permutation test results assessing the statistical significance of the first (LC1) and second (LC2) latent components, respectively. Histograms represent the null distributions of singular values obtained from 1000 permuted datasets in which the correspondence between BART and CPT-3 variables was randomly shuffled. The red vertical line indicates the observed singular value from the original (non-permuted) data. **Panels B and C** display the saliences of CPT-3 (A) and BART (B) variables on the first latent component (LC1). **Panels F and G** display the saliences of CPT-3 (D) and BART (E) variables on the second latent component (LC2). Bar height represents the salience of each variable, reflecting its contribution to the corresponding latent dimension extracted by PLS. Upward and downward bars indicate positive and negative contributions, respectively, defining the structure of each component. Dots represent salience distributions obtained from 500 bootstrap resamples. Variables highlighted in yellow show stable contributions, defined by 95% bootstrap confidence intervals that do not cross zero. **Panels D and H:** Scatterplots depicting the correspondence between BART- and CPT-3–derived subject scores for the first and second latent components, respectively. Each point represents a participant and is positioned according to the pair of component scores obtained by projecting that participant’s BART variables and CPT-3 variables onto the same latent component. The solid line denotes the least-squares regression fit. The Pearson correlation coefficient (r) and associated p-value quantify the strength and statistical significance of the between-domain association for each component.

On the D-BART side, LC1 was characterized by increased risk-taking (positive loadings on balloon explosion rate) and reduced overall task performance (negative loading on mean total points). Dynamic RT variables contributed more strongly and highlighted a response pattern characterized by RTs that were not only faster (negative loadings on mean-RT, BI-intercept, and DPC-intercept), but also less variable (negative loading on RT-standard-deviation), reflecting insufficient modulation of RT by both objective risk (negative loading on BI slope) and subjective uncertainty (positive loading on DPC slope).Importantly, this multivariate response pattern, characterized by combined flattening of DPC and BI RT slopes, mirrors the response-style differences that distinguished healthy controls from clinical participants in mixed-effects analyses and directly links these dynamics to maladaptive decisional outcomes. The strong correspondence between this D-BART pattern and the CPT-3 impulsivity signature supports the external validity of dynamic RT-based metrics as indicators of neurocognitive impulsivity captured by laboratory-based assessment.

PLS analysis also identified a second latent component (LC2) in which CPT-3 and D-BART response patterns were correlated across subjects (*r* = 0.42, *p* < .0001, Figure-2-Panel-H). Although this component accounted for a lower proportion of shared variance (13%), which did not exceed chance levels (*p* = .253, Figure-2-Panel-E), we report it here given its conceptual relevance. On the CPT-3 side, LC2 reflected a less conservative response style, with positive loadings on commission errors and negative loadings on omissions, combined with fast reaction times and pronounced RT slowing across task blocks, suggesting early impulsive responding followed by compensatory slowing (Figure-2-Panel-F). On the D-BART side, LC2 was characterized by slower RTs under low-risk conditions (positive BI intercept), reduced overall RT variability, and insufficient RT modulation by both objective and subjective risk (negative BI slope; positive DPC slope). This response style was again associated with reduced total points but, unlike LC1, was not driven by excessive risk-taking. Instead, it reflected premature cash-out under low-risk conditions, as indicated by negative loadings on balloon explosions and on pumps during successful trials (Figure-2-Panel-G). Thus, LC2 appears to capture a qualitatively distinct behavioral expression of insufficient cognitive control, impulsive risk avoidance rather than impulsive risk-taking. Despite opposite behavioral manifestations, both LC1 and LC2 share a common deficit in dynamically integrating contextual risk information into decision-making, and both are associated with impaired overall performance.

### Evaluating the Diagnostic and Clinical Relevance of BART-Derived Response Patterns

Having demonstrated the existence of a shared cognitive-control construct linking BART and CPT-3 response patterns, we next investigated whether the subject-specific readouts of cognitive control provided by the two instruments also predicted differential vulnerability to real-world behavioral manifestations of cognitive-control deficits. To this end, we multiplied participant-specific BART and CPT-3 behavioral patterns by the respective LC1 loadings derived from the population-level PLS analysis, yielding a pair of multivariate BART and CPT-3 scores for each participant. These scores, which captured the extent to which the LC1 cognitive-control pattern identified by the two instruments was expressed at the individual level, were then correlated with the severity of real-world behavioral symptoms of cognitive-control deficits, measured using the gold-standard parent-report Conners Rating Scales (CRS). Six symptom domains were considered: Inattention, Hyperactivity/Impulsivity, Learning Problems, Executive Dysfunction, Aggressiveness, and Peer Relationship Problems. Given emerging evidence suggesting gender-specific patterns in impulsivity expression deficits[59-63], we additionally tested whether gender moderates the association between BART/CPT-3 scores and the different CRS symptom dimensions.

BART-LC1 impulsivity scores were significantly correlated with Conners Hyperactivity/Impulsivity T-scores (r = 0.37, p = 0.007, Figure-3-Panel-A), whereas associations with other Conners subdomains were not statistically significant (Table 3). The association between BART-LC1 impulsivity and Hyperactivity/Impulsivity did not differ significantly between males and females (Gender × BART interaction: β=1.57, p=0.52, R-males=0.44, P-Males=0.03, R-females=0.28, p-females=0.15). However, a significant Gender × BART interaction was observed for Peer Relationship Problems (β=-5.9, p=0.01), indicating a female-specific association between BART-LC1 and Peer Relationships that was not observed in males (R-Female=0.45, P-Female=0.02, R-Male=-0.22, P-Male=0.31, Figure-3-Panel-B). To capture such combined gender-specific effects we computed a gender-specific clinical redout composed of exclusively of Hyperactivity/Impulsivity in males and the average of Hyperactivity/Impulsivity and Peer Relationship Problems in females. The resulting score was strongly correlated with BART-LC1 (r = 0.45, p = 0.001, Figure-3-Panel-C). Population specific factors did not significantly drive this association which remained significant when restricting the analysis to exclude HC (R=0.39, p=0.009) as well as when retaining only the ADHD clinical subpopulation (R=0.41, p=0.011).

**Table 1.** Demographic and assessment characteristics by group.

| Sample characteristics | Population | N | Male | Female | Mean Age | SD Age | Mean Number of BART Assessments | Subject with CPT | Subject with Conners |
| --- | --- | --- | --- | --- | --- | --- | --- | --- | --- |
|  | 22q | 10 | 4 | 6 | 16,66 | 2,7 | 2,8 | 10 | 8 |
|  | ADHD | 40 | 18 | 22 | 15,43 | 1,9 | 32,3 | 40 | 37 |
|  | HC | 9 | 6 | 3 | 16,76 | 3,65 | 2,7 | 9 | 6 |
| Between-group Comparisons | Population Comparison | $\chi^2$<br>(Sex Distribution) | p<br>(Sex Distribution) | t (Age) | p (Age) | t<br>(Number of BART Assessments) | p<br>(Number of BART Assessments) | | |
|  | 22q vs ADHD | 0,08 | 0,78 | 1,66 | 0,10 | -3,68 | 0,00 |  |  |
|  | 22q vs HC | 1,35 | 0,25 | -0,07 | 0,95 | 0,35 | 0,73 |  |  |
|  | ADHD vs HC | 1,38 | 0,24 | -1,56 | 0,13 | 3,51 | 0,00 |  |  |

Sample size (N), sex distribution (male/female), mean age ( ± SD), mean number of ecological BART assessments per participant (N\_BART), and number of available CPT and Conners assessments are reported separately for the 22q11DS, ADHD, and healthy control (HC) groups.
Between-group differences in age were assessed using independent-samples *t*-tests. Differences in sex distribution were evaluated using chi-square ( $\chi^2$ ) tests. Corresponding test statistics and p-values are reported in the table.

**Table 2.**
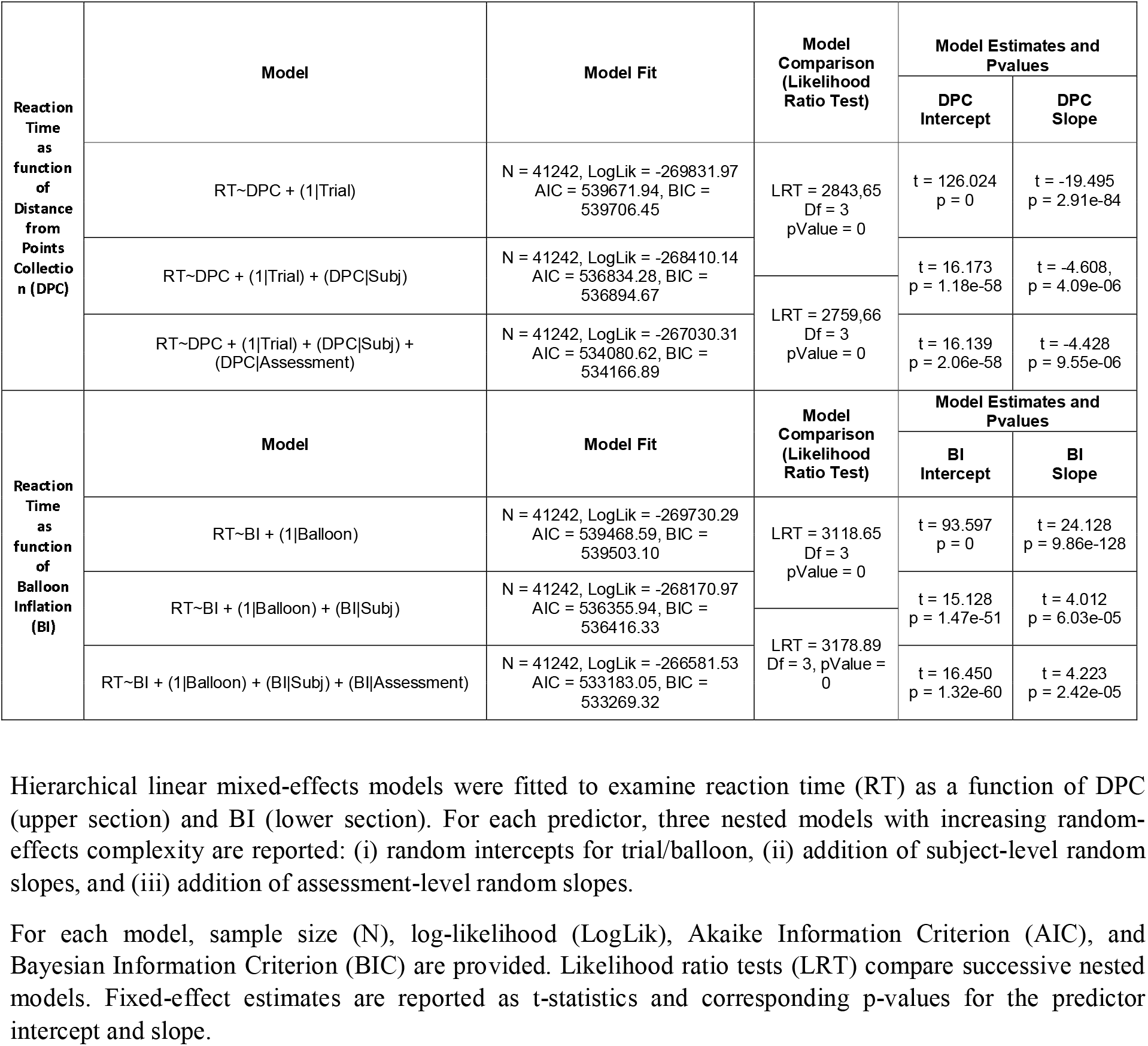
Hierarchical linear mixed-effects models of reaction time as a function of distance from point collection (DPC) and balloon inflation (BI).

**Table 3.** Associations between participant-level scores on the first CPT–BART PLS component and Conners symptom scales.

| D-BART-1 | CONNERS SCALE | All Sample | Male | Female | Interaction Model: $\text{Conners} \sim 1 + \text{D-BART-1} + \text{D-BART-1} * \text{Gender}$ |
| --- | --- | --- | --- | --- | --- |
| | Inattention | $r = 0.14$<br>$p = 0.32$ | $r = 0.08$<br>$p = 0.73$ | $r = 0.20$<br>$p = 0.30$ | D-BART-1*gender = -1.19; $p = 0.61$; $R^2 = 0.03$<br>D-BART-1 = 1.76; $p = 0.30$; gender = 0.08; $p = 0.9$ |
| | Hyperactivity | **$r = 0.37$**<br>**$p = 0.01$** | $r = 0.44$<br>$p = 0.03$ | $r = 0.28$<br>$p = 0.15$ | D-BART-1*gender = 1.58; $p = 0.52$; $R^2 = 0.20$<br>D-BART-1 = 2.45; $p = 0.17$; gender = 8.11; $p = 0.08$ |
| | Learning Problems | $r = 0.07$<br>$p = 0.64$ | $r = 0.06$<br>$p = 0.80$ | $r = 0.07$<br>$p = 0.73$ | D-BART-1*gender = -0.10; $p = 0.97$; $R^2 = 0.01$<br>D-BART-1 = 0.55; $p = 0.75$; gender = 1.68; $p = 0.70$ |
| | Executive Functions | $r = 0.04$<br>$p = 0.77$ | $r = -0.02$<br>$p = 0.93$ | $r = 0.09$<br>$p = 0.66$ | D-BART-1*gender = -0.78; $p = 0.69$; $R^2 = 0.01$<br>D-BART-1 = 0.66; $p = 0.64$; gender = 1.10; $p = 0.76$ |
| | Aggressiveness | $r = 0.12$<br>$p = 0.41$ | $r = -0.05$<br>$p = 0.82$ | $r = 0.28$<br>$p = 0.16$ | D-BART-1*gender = -2.97; $p = 0.21$; $R^2 = 0.07$<br>D-BART-1 = 2.62; $p = 0.13$; gender = -4.62; $p = 0.29$ |
| | Peer Relationship | $r = 0.12$<br>$p = 0.39$ | $r = -0.22$<br>$p = 0.31$ | **$r = 0.45$**<br>**$p = 0.02$** | **D-BART-1\*gender = -5.90; $p = 0.01$**; $R^2 = 0.16$<br>D-BART-1 = 4.23; $p = 0.01$; gender = -5.59; $p = 0.19$ |
| | HyPr | **$r = 0.45$**<br>**$p = 0.001$** | $r = 0.44$<br>$p = 0.034$ | **$r = 0.45$**<br>**$p = 0.015$** | D-BART-1*gender = 0.69; $p = 0.75$; $R^2 = 0.26$<br>D-BART-1 = 3.34; $p = 0.04$; gender = 7.33; $p = 0.07$ |
| D-CPT-1 | CONNERS SCALE | All Sample | Male | Female | Interaction Model: $\text{Conners} \sim 1 + \text{D-CPT-1} + \text{D-CPT-1} * \text{Gender}$ |
| | Inattention | $r = 0.12$<br>$p = 0.39$ | $r = 0.04$<br>$p = 0.84$ | $r = 0.19$<br>$p = 0.32$ | D-CPT-1*gender = -1.11; $p = 0.62$; $R^2 = 0.02$<br>D-CPT-1 = 1.45; $p = 0.32$; gender = 0.92; $p = 0.83$ |
| | Hyperactivity | $r = 0.18$<br>$p = 0.20$ | $r = 0.13$<br>$p = 0.55$ | $r = 0.32$<br>$p = 0.10$ | D-CPT-1*gender = -1.21; $p = 0.62$; $R^2 = 0.12$<br>D-CPT-1 = 2.46; $p = 0.13$; gender = 9.94; $p = 0.04$ |
| | Learning Problems | $r = 0.18$<br>$p = 0.21$ | $r = 0.12$<br>$p = 0.57$ | $r = 0.24$<br>$p = 0.21$ | D-CPT-1*gender = -0.71; $p = 0.75$; $R^2 = 0.04$<br>D-CPT-1 = 1.72; $p = 0.24$; gender = 2.54; $p = 0.55$ |
| | Executive Functions | $r = 0.07$<br>$p = 0.64$ | $r = 0.04$<br>$p = 0.84$ | $r = 0.09$<br>$p = 0.63$ | D-CPT-1*gender = -0.35; $p = 0.85$; $R^2 = 0.01$<br>D-CPT-1 = 0.61; $p = 0.62$; gender = 1.44; $p = 0.69$ |
| | Aggressiveness | $r = 0.08$<br>$p = 0.58$ | $r = -0.16$<br>$p = 0.47$ | $r = 0.21$<br>$p = 0.28$ | D-CPT-1*gender = -2.89; $p = 0.20$; $R^2 = 0.06$<br>D-CPT-1 = 1.75; $p = 0.24$; gender = -4.05; $p = 0.36$ |
| | Peer Relationship | $r = 0.14$<br>$p = 0.32$ | $r = -0.01$<br>$p = 0.96$ | $r = 0.22$<br>$p = 0.26$ | D-CPT-1*gender = -1.89; $p = 0.42$; $R^2 = 0.05$<br>D-CPT-1 = 1.81; $p = 0.24$; gender = -4.51; $p = 0.32$ |
| | HyPr | $r = 0.18$<br>$p = 0.21$ | $r = 0.13$<br>$p = 0.55$ | $r = 0.33$<br>$p = 0.08$ | D-CPT-1*gender = -0.89; $p = 0.69$; $R^2 = 0.12$<br>D-CPT-1 = 2.14; $p = 0.15$; gender = 9.26; $p = 0.04$ |

Pearson correlation coefficients ( $r$ ) and corresponding $p$ -values are reported between first-dimension CPT scores (D-CPT-1; lower section) and first-dimension BART scores (D-BART-1; upper section) and Conners subscales (Inattention, Hyperactivity, Learning Problems, Executive Functions, Aggressiveness, Peer Relationship Problems and HyPr), computed for the full sample and separately for males and females.
To test for sex moderation, linear regression models of the form $\text{Conners scale} \sim 1 + \text{D-CPT-1} + \text{D-CPT-1} \times \text{gender}$ (upper section) and $\text{Conners scale} \sim 1 + \text{D-BART-1} + \text{D-BART-1} \times \text{gender}$ (lower section) were fitted for each subscale. For each model, test statistics and $p$ -values for the first-dimension score, gender main effect, and interaction term are reported, together with model $R^2$ .

**Table 4.**
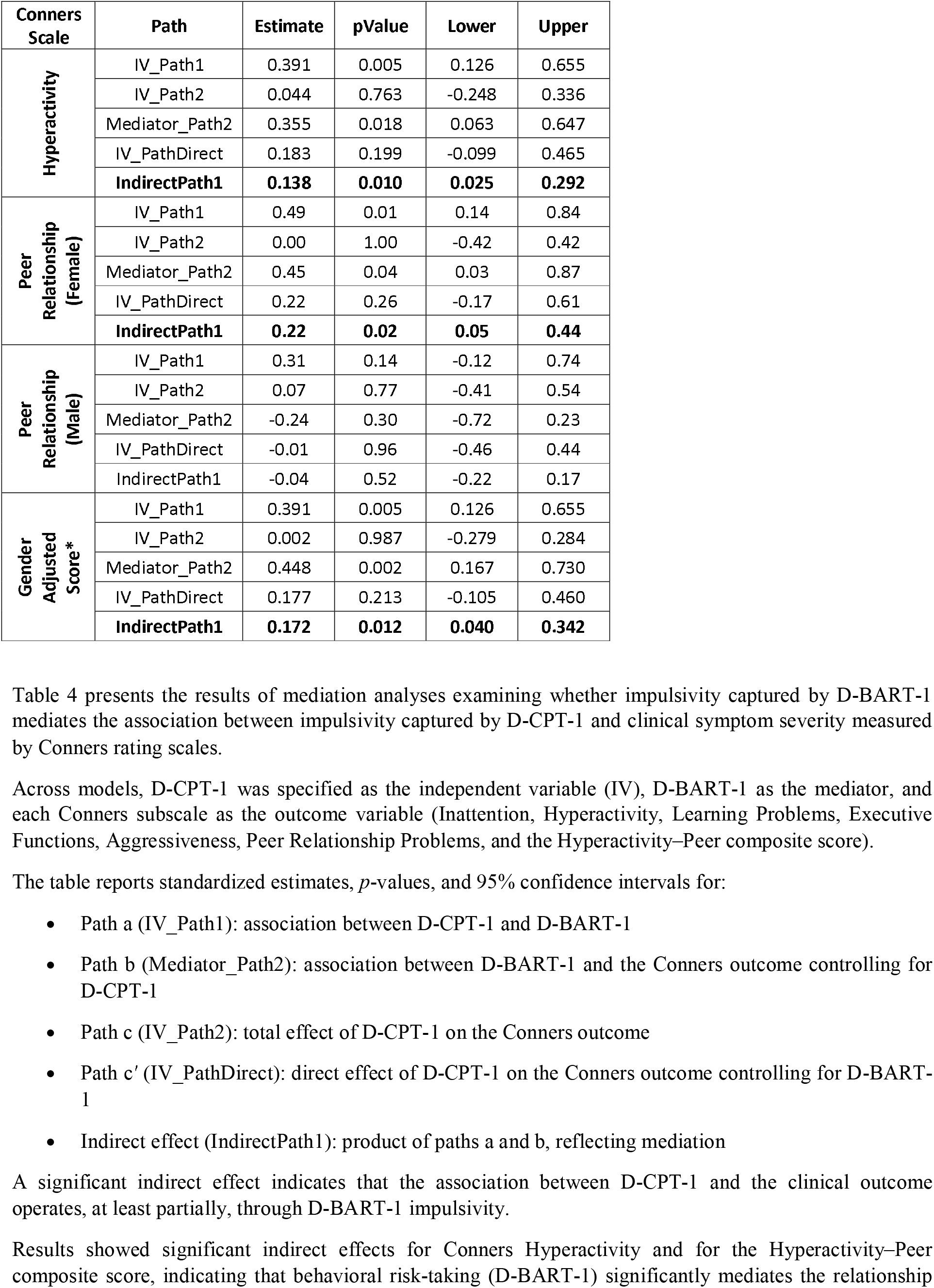
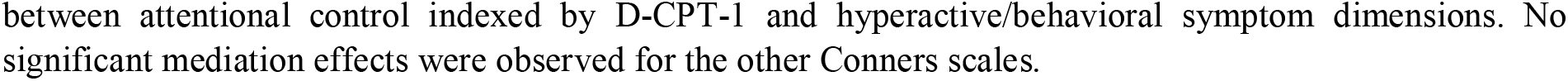
Mediation Analysis.

**Figure 3:**
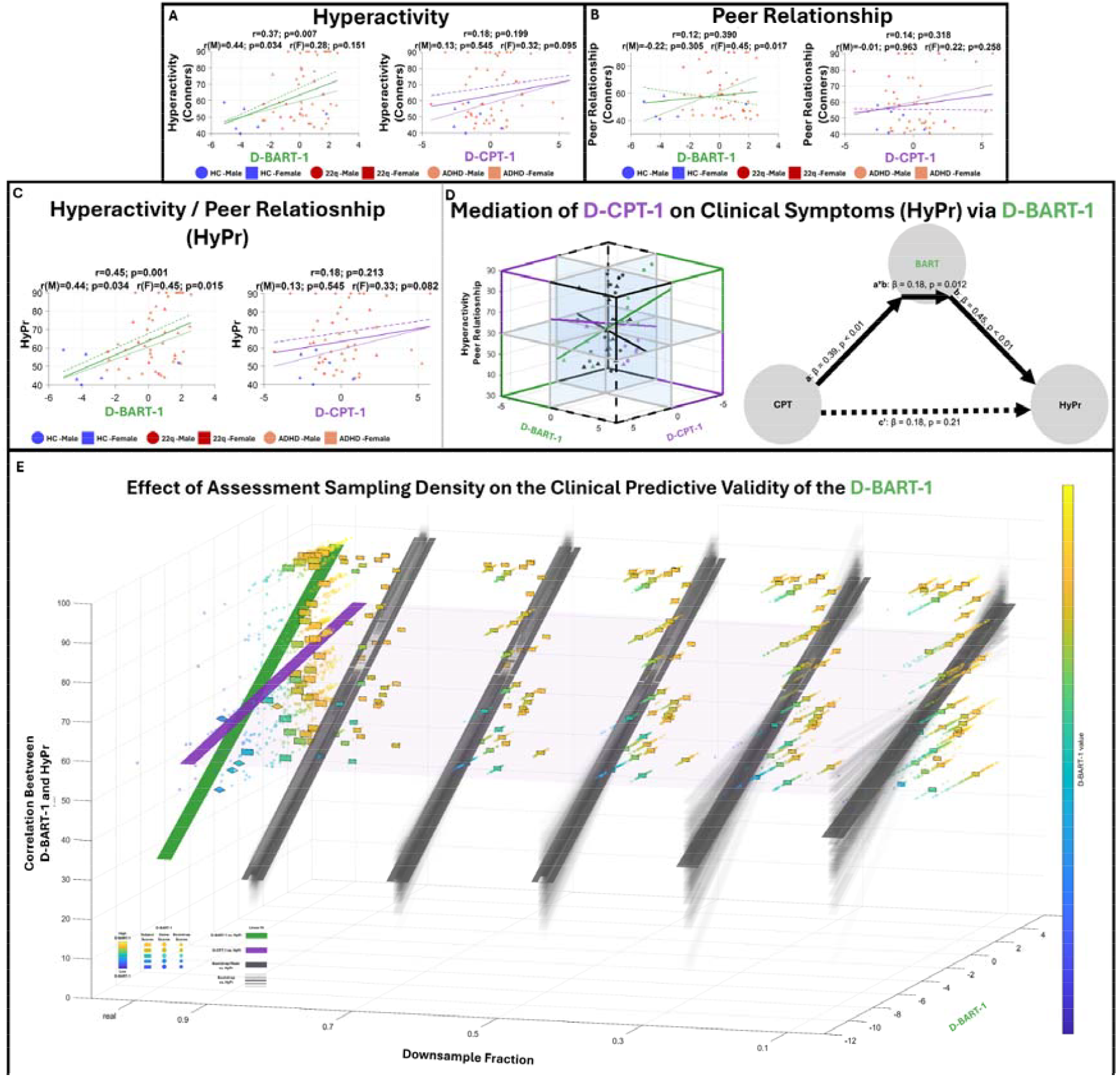
Panels A, B, and C. Associations between LC1 PLS scores and Conners symptom scales. Scatterplots show correlations between LC1 PLS scores derived from BART (Fig. 1 within each panel) and CPT (Fig. 2 within each panel) and Hyperactivity/Impulsivity (Panel A), Peer Relationship Problems (Panel B), and a sex-specific composite score, defined as Hyperactivity/Impulsivity in males and the mean of Hyperactivity/Impulsivity and Peer Relationship Problems in females. (Panel C). Points denote individuals (blue: healthy controls; red: 22q11DS; orange: ADHD; circles: males; triangles: females). Regression lines are shown for BART (green) and CPT (violet); solid lines indicate the full sample, dashed males, and dotted females. Pearson correlation coefficients and two-sided p-values are reported within each subpanel. **Panel D-1. Three-dimensional association between task-derived dimensions and clinical symptom severity**. Three-dimensional scatter plot depicting the relationship between first-dimension BART scores (D-BART-1; x-axis), first-dimension CPT scores (D-CPT-1; y-axis), and the composite Hyperactivity/Impulsivity score (HyPr; z-axis). Planes intersect each axis at predefined thresholds: HyPr at the clinical cut-off (≥ 60), and D- BART-1 and D-CPT-1 at their respective optimal thresholds derived from receiver operating characteristic (ROC) analyses predicting clinical HyPr status. Plot borders are color-coded according to classification agreement relative to the HyPr clinical cut-off. Black borders indicate regions in which both D-BART-1 and D-CPT-1 classify diagnosis concordantly; solid lines denote correct classification, whereas dashed lines denote incorrect classification. Green borders indicate regions in which D-BART-1 classifies diagnosis correctly while D-CPT-1 does not; violet borders indicate regions in which D-CPT-1 classifies correctly while D-BART-1 does not. Regression lines are overlaid for the associations between D-BART-1 and D-CPT-1 (black), D-BART-1 and HyPr (green), and D-CPT-1 and HyPr (violet). **Panel D-2. Mediation model linking CPT-, BART-, and symptom-derived first-dimension scores**. Path diagram illustrating a mediation model in which first-dimension CPT scores (D-CPT-1) predict the composite symptom score (HyPr) directly and indirectly via first-dimension BART scores (D-BART-1). The effect of CPT on BART is shown on the arrow from CPT to BART (*a*). The effect of BART on HyPr, estimated while controlling for CPT, is shown on the arrow from BART to HyPr (*b*). The direct effect of CPT on HyPr, estimated while controlling for BART, is shown on the arrow from CPT to HyPr (*c*′). The indirect (mediated) effect is quantified as the product of these two paths (*a × b*). Standardized regression coefficients (β) and corresponding p-values are displayed for each structural path and for the indirect effect. Solid arrows denote statistically significant effects; dashed arrows indicate non-significant effects. **Panel E. Effects of down sampling D-BART-1 data ability on symptom severity prediction accuracy**. Three-dimensional visualization of the relationship between subject-level D-BART-1 and composite Hyperactivity/Impulsivity–Peer Relationship Problems scores across progressive reductions in data availability. Subject-level mean D-BART-1 and composite symptom scores are represented along the X- and Y-axes, respectively. The Z-axis represents the impact of downsampling the proportion of retained BART observations in the ADHD group across progressively more restrictive thresholds (fractions: 1, 0.9, 0.7, 0.5, 0.3, 0.1). The first plane along the Z-axis corresponds to the full dataset (downsampling fraction = 1). Vertical subject markers indicate observed subject-level mean D-BART-1 values, with ADHD and healthy control (HC) participants distinguished by different marker geometries. Scattered points represent single-game D-BART-1 observations from the original dataset. The green surface denotes the regression fit between subject-level mean D-BART-1 and Hyperactivity/Impulsivity–Peer Relationship Problems scores across all participants, whereas the violet surface represents the corresponding fit for CPT scores. Larger semi-transparent planes extend these real-data fits into the downsampling space to facilitate comparison with bootstrap-derived associations. For each downsampling threshold along the Z-axis, 1,000 random subsampling iterations were performed, of which 100 representative iterations are shown for visualization. Scatter points represent variation in subject-specific D-BART-1 values across iterations, while rectangular surfaces represent the average subject-specific D-BART-1 value across all iterations. Dark gray ribbons indicate the mean regression slope across all iterations at a given threshold, whereas semi-transparent gray ribbons represent the distribution of iteration-specific regression slopes around this mean, with darker ribbons corresponding to fits closer to the average regression across bootstrap samples. Comparing regression distributions across downsampling thresholds provides a quantitative representation of how reduced D-BART data availability impacts the accuracy with which D-BART captures between-subject variability in Hyperactivity/Impulsivity–Peer Relationship Problems severity scores.

Despite significant associations between BART-LC1 and CPT-LC1 impulsivity scores (r = 0.37, p = 0.0037), CPT-LC1 scores were not significantly correlated with any individual Conners subdomain (Table 3), nor with the gender-informed composite score combining Hyperactivity/Impulsivity and Peer Relationship Problems (r=0.18, p=0.213). Indeed, BART-LC1 scores significantly mediated the association between CPT-LC1 and Conners Hyperactivity/Impulsivity scores (β=0.138, p=0.01), as well as the female-specific association with Peer Relationship Problems (β=0.22, p=0.02) and with the composite Hyperactivity/Impulsivity/Peer-Relationship-Problems score (β=0.172, p=0.012, Figure-3-Panel-D). These findings indicate that although CPT-3 and BART capture overlapping variance in neurocognitive impulse-control performance, the impulsivity signal derived from the ecologically administered BART is more closely linked to clinically meaningful real-life behavioral manifestations of impulse-control deficits.

Finally, we examined whether the repeated ecological assessment framework of the BART, allowing dynamic fluctuations in cognitive control to be captured across multiple days and contexts, contributed to its superior clinical performance relative to the single-session laboratory-based CPT-3. To isolate the added value of repeated assessments, we derived assessment-specific cognitive-control indices by projecting BART-LC1 loadings onto assessment-specific BART metrics. We then randomly downsampled the proportion of available assessments 1,000 times at different availability thresholds (90%, 70%, 50%, 30%, 10%). At each iteration, BART-LC1 scores were averaged across the retained assessments for each participant and correlated with the Conners Hyperactivity/Impulsivity–Peer Relationship Problems composite score. By comparing the distributions of correlation coefficients across availability thresholds, we quantified the impact of reduced sampling density on the ability of BART to capture between-subject variation in symptom severity.

Results revealed a highly significant, progressive decline in the average correlation strength between BART and symptom severity as data availability decreased (F = 389.65, p<.0001), accompanied by increased variability in estimation (Figure-3-Panel-E). These findings demonstrate that the superior accuracy of BART in capturing behavioral symptom severity is directly related to the ecological framework in which it was administered, with diagnostic performance tightly linked to the precision with which dynamic cognitive-control fluctuations are measured across repeated assessments.

### Discussion

Impulse-control deficits have major societal consequences[12, 64, 65], yet existing neurocognitive paradigms have limited ability to predict clinically meaningful behavioral outcomes[36, 66]. Here, we show that combining repeated ecological assessment with computational modelling of response-time (RT) dynamics substantially improves the characterization of impulse control.

Our modelling framework was anchored in dual-process theories of decision-making, which propose that increasing risk and uncertainty shift behavior from fast, automatic responding toward slower, deliberative control[51, 54]. While richly informative, these theories have previously been validated using highly parameterized frameworks that explicitly model interacting cognitive systems [51, 52, 56, 67], limiting their clinical applicability and suitability for repeated ecological assessment. Our first objective was therefore to test whether the central predictions of dual-process theories could be meaningfully examined using a more parsimonious modelling framework. Specifically, RT was modelled as a function of balloon inflation (objective risk) and distance from point collection (DPC), a proxy for subjective uncertainty, while a mixed-effects framework characterized both subject-level and assessment-level variability in response style. In accordance with dual-process theories, we found that RT increased with balloon inflation and peaked near cash-out decisions. Moreover, subject-specific random effects significantly improved model fit, indicating that individuals differ in their tendency to shift from fast to deliberative processing as risk and uncertainty increase—a potential mechanism underlying vulnerability to maladaptive impulsive decisions [49, 50]. Interestingly, assessment-specific random effects further improved model fit, supporting the notion that impulse control is not solely a stable trait but also fluctuates with changing states and contexts, including sleep, arousal, and affect [40, 43, 44].

Next, we validated our modelling framework by testing its clinical relevance in differentiating healthy controls from clinical participants. Compared with clinical participants, healthy controls showed greater RT slowing with increasing objective risk and subjective uncertainty, whereas baseline RT differed little between groups. These findings suggest that clinical participants were not simply faster responders but were less able to recruit deliberative control as contextual demands increased. Although greater RT slowing with increasing objective risk could alternatively reflect lower risk tolerance rather than stronger cognitive control [53], modelling subjective uncertainty produced a similar pattern, supporting the interpretation that impaired recruitment of deliberative control distinguishes clinical participants from healthy controls. Of note, conventional BART performance metrics failed to differentiate clinical participants from healthy controls, highlighting the added value of dynamic RT modelling.

Our next objective was to determine whether dynamic RT modelling could capture a common cognitive-control dimension bridging neurocognitive paradigms and traditional performance metrics. Our multivariate analysis identified a primary BART latent component characterized by reduced RT adaptation to both objective risk and subjective uncertainty and associated with poorer task performance, including more balloon explosions and fewer points earned. Importantly, this pattern covaried with a CPT-3 profile characterized by impaired target discrimination and excessively rapid responding. Although the BART and CPT are traditionally viewed as measuring higher- and lower-order forms of impulsivity, respectively, our findings suggest that both tasks capture a common deficit in adaptive cognitive control, expressed as inadequate modulation of response strategies according to contextual demands. This interpretation aligns with broader conceptions of cognitive control as the capacity to flexibly regulate speed–accuracy trade-offs in response to changing contextual demands [49]. Within this framework, performance-based indices of inhibitory failure across paradigms may represent downstream manifestations of a shared, transdiagnostic deficit in adaptive cognitive control that becomes more apparent when task outcomes and response-style dynamics are jointly modelled using multivariate approaches [49].

Our results also suggest that, when considered in isolation, traditional performance metrics such as proportion of exploded balloons, summarize behavioral outcomes but may fail to distinguish the cognitive mechanisms that generate those outcomes. Elevated risk-taking may reflect greater risk tolerance rather than impaired cognitive control, whereas maladaptive cognitive control may equally manifest as excessive risk avoidance under low-risk conditions [53]. Consistent with this interpretation, LC2 identified a qualitatively distinct behavioral profile characterized by reduced adaptation of RT to contextual risk together with premature and maladaptive risk avoidance under low-risk conditions. This pattern supports the notion that impaired top-down control contributes to dysregulated responses to both appetitive and aversive signals [81], potentially helping explain the frequent co-occurrence of impulsivity, emotional dysregulation, and anxiety [82–86]. The ability to dissociate common mechanisms underlying seemingly opposite behavioral phenotypes may provide a useful framework for understanding clinical heterogeneity [87,88]. More broadly, our multivariate analytic approach revealed a shared neurocognitive construct that is consistent with genetic evidence for common biological liability across disorders characterized by impaired impulse control[12, 68]. This shared construct may also help reconcile evidence for a common impulsivity dimension with the poor external validity of existing neurocognitive paradigms [25, 32, 69]. Our findings suggest that this discrepancy arises less from the paradigms themselves than from the conventional performance metrics used to quantify them.

Our final objective was to evaluated if the combination of ecological neurocognitive testing and advanced analytics could be translated into substantially greater clinical relevance compared to conventional laboratory assessment. We found that, although D-BART and CPT-3 captured a shared latent dimension of cognitive control, only ecologically derived D-BART measures were directly associated with real-world behavioral impulsivity. Mediation analyses showed that the association between CPT-3 performance and behavioral symptom severity was fully mediated by D-BART measures, suggesting that the clinically relevant variance captured by the CPT-3 is largely explained by the cognitive-control processes measured through repeated ecological assessment. Together, these findings indicate that repeated ecological assessment does not simply provide an alternative measure of impulse control but captures clinically relevant fluctuations in cognitive-control processes that remain largely inaccessible to conventional laboratory paradigms. To our knowledge, this is the first direct demonstration that an ecologically administered neurocognitive paradigm provides incremental diagnostic value over an established laboratory gold standard. Our final analyses provide important insight into the source of this improved clinical performance. Rather than reflecting intrinsic differences between the BART and CPT paradigms, the advantage appeared to arise primarily from repeated ecological assessment. Cognitive-control estimates varied substantially across repeated assessments, and progressively reducing the number of observations markedly weakened associations with behavioral symptoms. In other words, degrading the precision with which dynamic fluctuations in cognitive control were sampled directly reduced the ability to identify clinically meaningful individual differences. These findings support an emerging view that impulsivity is better conceptualized as a dynamic distribution of cognitive-control states across changing contexts than as a fixed trait measured during a single laboratory session [40, 43, 44, 70]. More broadly, they provide direct empirical support for ecological neurocognitive assessment as a means of bridging the longstanding gap between laboratory measures and real-world behavior[71, 72].

Several limitations should be acknowledged. Ecological monitoring protocols differed across participant groups, and 22q11.2 deletion syndrome represents a broader psychiatric phenotype than ADHD alone[73]. Nevertheless, the principal findings remained significant when analyses were restricted to the ADHD sample, suggesting that they were not driven by population-specific characteristics.

Despite these limitations, the study has several important strengths. To our knowledge, this is the first study to demonstrate that repeated ecological neurocognitive assessment, combined with computational modelling of RT dynamics, provides greater clinical utility than a gold-standard laboratory paradigm. More broadly, our findings suggest that the longstanding gap between laboratory measures of impulsivity and clinically meaningful behavioral outcomes may reflect the outcome metrics traditionally extracted from neurocognitive paradigms rather than limitations of the paradigms themselves. By combining repeated ecological assessment with computational modelling, existing paradigms can instead be used to isolate latent cognitive-control processes that generalize across tasks and more closely predict real-world behavioral vulnerability. Given the substantial individual and societal burden associated with impaired impulse control[12, 64, 65], improving early identification of vulnerable individuals represents an important public health priority[23]. This need may become increasingly pressing as contemporary environments expose individuals to an expanding array of immediately rewarding stimuli [74-80] may place greater demands on impulse control and thus disproportionately affect individuals with impaired self-regulation[81-86] If replicated, the proposed smartphone-based framework could provide a scalable approach for identifying at-risk individuals before adverse behavioral consequences become established, thereby facilitating earlier and more targeted preventive interventions.

## Methods

### Participants and Study Design

Sixty adolescents and young adults participated in the study: 40 individuals with recently diagnosed idiopathic ADHD (18M/22F; mean age = 15.43 years, SD = 1.90) recruited through the Platform for the Evaluation of Executive Functions at the Fondation Pôle Autisme (Geneva, Switzerland); 10 individuals with 22q11.2 deletion syndrome (22q11DS; 4M/6F; mean age = 16.66 years, SD = 2.70) recruited through the 22q11DS Swiss longitudinal cohort at the University of Geneva [87, 88]; and 9 healthy controls (HC; 6M/3F; mean age = 16.76 years, SD = 3.65), recruited among unaffected siblings of 22q11DS participants. HC exclusion criteria included premature birth and any lifetime history of psychiatric, neurological, or learning disorders.

All participants were aged between 12 and 30 years and had sufficient proficiency in French to complete the assessments. Written informed consent was obtained from all participants or their legal guardians, as appropriate. The study protocol was approved by the Swiss Ethics Committees on Human Research (CCER).

### Clinical and Laboratory-Based Neurocognitive Assessment

DSM-5 diagnostic criteria for ADHD [89], were evaluated through structured clinical interviews (*Schedule for Affective Disorders and Schizophrenia for School-Age Children-Present and Lifetime Version (K-SADS-PL*)[90] by trained clinicians in both ADHD and 22q11DS clinical populations. ADHD Behavioral symptom severity was assessed using the Conners Rating Scales (CRS), (CRS) [91], with age- and sex-normalized T-scores analyzed for six subscales: Inattention, Hyperactivity/Impulsivity, Learning Problems, Executive Dysfunction, Aggressiveness, and Peer Relationship Problems. Parent-report CRS served as the primary behavioral outcomes (available for 37 participants).

All participants underwent a comprehensive neurocognitive assessment battery comprising the Conners Continuous Performance Test, Third Edition (CPT-3) [92], the Continuous Auditory Test of Attention (CATA) [93], the Tower of London task [94], the Trail Making Test [95, 96], and working memory subtests from the Wechsler Intelligence Scales [97, 98].

The present study focused on CPT-3 variables that index attentional control and impulsive responding. Specifically we analyzed eight age-normed T-score variables: Detectability (d′), reflecting the ability to discriminate between target and non-target stimuli; Omissions, indicating failures to respond to target stimuli; Commissions, reflecting incorrect responses to non-target stimuli; Perseverations, defined as extremely rapid responses (<100 ms); Hit Reaction Time (RT), representing the mean response speed for non-perseverative responses; Hit RT Standard Deviation, indexing variability in reaction time across the task; Variability, a within-subject measure of response speed consistency across task segments; Hit RT Block Change, capturing changes in reaction time over the duration of the task (with slowing indicating declining sustained attention); and Hit RT Inter-Stimulus Interval (ISI) Change, reflecting changes in reaction time across varying stimulus intervals, with slower responses at longer intervals suggesting reduced vigilance.

### Smartphone-Based Ecological Neurocognitive Assessment

Ecological assessments were delivered via the LAMP digital phenotyping platform [99]. The smartphone battery included digital Trail Making Test A and B[95], a Spatial Working Memory task, and a smartphone implementation of the Balloon Analogue Risk Task (BART)[31]. Participants received notifications three times per day prompting completion of the neurocognitive battery. The order of task presentation was randomized at each administration to control for potential fatigue and order effects.

The 22q11DS and HC groups completed the smartphone protocol over two days on a standardized device provided by the study team (Xiaomi Redmi Note 12S 4G LTE), in parallel with their in-person evaluation in Geneva. The ADHD group completed a one-month ecological protocol on their personal smartphones, embedded within a broader digital phenotyping design that also included EMA surveys (eight per day), a morning sleep survey, an evening symptom/function survey, and passive data collection via wearable devices. Participants were reimbursed proportionally to engagement (50–250 CHF).

### Digital BART Task and Derived Measures

In the present paper we focused specifically on results of the Balloon Analogue Risk Task designed to capture neurocognitive impulsivity. In this task, participants inflated a virtual balloon, earning one point per pump while risking balloon explosion and loss of accumulated points. Participants could choose to “cash out” at any time, balancing risk and reward. Each session consisted of 10 balloons. While inspired by the original BART developed by Lejuez et al. (2002), our version introduces several key structural modifications. Unlike the original task, which features an average explosion point of approximately 64 pumps (SD = 32), our implementation employed a substantially lower explosion threshold (mean = 10 pumps, SD = 5), limited each session to 10 balloons, and was administered repeatedly across multiple days. These design choices facilitated brief, repeated assessments, supporting ecologically valid, high-frequency data collection without overburdening participants.

To ensure data quality and meaningful estimation of behavioral indices, several exclusion criteria were applied to the BART data The first game completed by each participant was removed a priori because that game served as a familiarization phase. We then retained only participants with at least two remaining games, corresponding to at least three completed games in total. After these initial selection steps, the dataset comprised 14 participants / 37 games / 1,557 clicks in the 22q11DS group, 40 / 1,327 / 66,108 in the ADHD group, and 9 / 24 / 1,079 in the healthy control group. Additional participant-level exclusion removed participants whose data did not contain both explosion and non-explosion outcomes. We next applied a game-level exclusion by removing games with fewer than 3 pumps, followed by click-level exclusions consisting of the first click of each balloon and RT outliers above the empirical 99.95th percentile of the RT distribution (RT > 3853.35 ms). After all exclusions, the final analytic dataset comprised 10 participants / 28 games / 1,137 clicks in the 22q11DS group, 40 / 1,295 / 52,773 in the ADHD group, and 9 / 24 / 837 in the healthy control group. The mean number of successfully completed game sessions per participant was 2.67 ± 0.71 in the HC group, 2.80 ± 0.92 in the 22q11DS group, and 32.38 ± 25.20 in the ADHD group.

Traditional BART metrics were computed at the session level and then aggregated within participant, including: mean collected points, proportion of exploded balloons, mean inflation at point collection (adjusted pumps), mean RT, and RT standard deviation. In addition, dynamic response-style parameters were derived from mixed-effects models of RT modulation by objective risk and subjective uncertainty (see below), and similarly aggregated within participant.

### Statistical Analysis

#### Mixed-Effects Modeling of RT Dynamics

We modeled response-time (RT) dynamics to quantify how decision speed was modulated by (i) objective risk, indexed by balloon inflation level (number of pumps; BI), and (ii) subjective uncertainty, indexed by distance from point collection (DPC), defined as the number of remaining pumps before the participant cashed out on successful balloons. RT corresponded to the time between successive pumps within a balloon (ClickDuration). Because multiple clicks were nested within balloons, balloons within sessions (games), and sessions within participants, we used linear mixed-effects models to account for the resulting dependency structure. RT analyses were restricted to successful balloons ending in point collection.

##### Objective risk model (RT as a function of balloon inflation)

We first estimated a baseline model capturing the population-level effect of objective risk while accounting for within-balloon dependencies via a balloon-level random intercept:

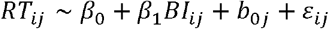

where *RT*_*ij*_ denotes the reaction time of click *i* within balloon *j, BI*_*ij*_ denotes the pump number (inflation level), *β*_*0*_is the fixed intercept, *β*_1_ is the fixed effect of balloon inflation (objective risk), *b*_*0j*_is the balloon-specific random intercept, and *ε*_*ji*_ is the residual error term.

##### Subjective uncertainty model (RT as a function of distance from point collection)

To quantify RT modulation by subjective uncertainty, we modeled RT as a function of DPC on successful balloons (cash-out trials). We first fit a balloon random-intercept model:

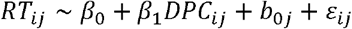

##### Between-subject variability in RT dynamics

Next, to test whether modulation of RT by risk and uncertainty varied across individuals, we extended these models to include participant-level random intercepts and random slopes:

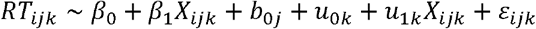

where *k*indexes participants and (*u*_*0k*_, *u*_*1k*_ )are participant-specific deviations in baseline RT and *X*-related sensitivity, with *X*_*ijk*_denoting either *BI*_*ijk*_or *DPC*_*ijk*_.

Likelihood ratio tests (LRTs) were used to compare nested models and evaluate the added explanatory value of participant-level random effects.

##### Within-subject variability across repeated assessments

Finally, to capture within-person fluctuations across repeated ecological assessments, we extended the model to include session-level (game-level) random intercepts and slopes:

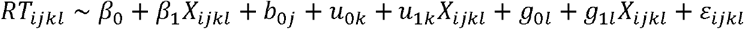

where *l* indexes sessions (games) and (*g*_*0i*_,*g*_*1i*_)capture session-specific deviations in baseline RT and *X*-related modulation, with *X*-denoting either *BI*_*ijk*_or *DPC*_*ijk*_.

Nested models were compared using likelihood ratio tests (LRTs) to evaluate the added explanatory value of session-level random effects.

##### Diagnostic effects on RT dynamics

To test whether RT dynamics differed by diagnostic group, we fit mixed-effects models including diagnosis (HC vs clinical participants [ADHD + 22q11DS]) and its interaction with BI or DPC:

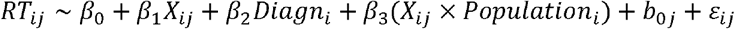

where *X*_*ij*_denotes either *BI*_*ij*_or *DPC*_*ij*_, and *β*_3_captures population (HC-CP) differences in risk- or uncertainty-related RT modulation.

##### Deriving participant-level response-style indices

To obtain participant-level dynamic response-style indices comparable to traditional BART metrics, we extracted session-level random-effect estimates from models including balloon-level random intercepts and session-level random intercepts and slopes (for BI and DPC). For each participant, we averaged session-specific estimates across sessions to derive four summary parameters:

- *RT-by-BI intercept and RT-by-BI slope*
- *RT-by-DPC intercept and RT-by-DPC slope*

For comparison, we also computed participant-level averages for traditional BART metrics: mean collected points, proportion of exploded balloons, mean inflation at point collection, mean RT, and RT standard deviation. These participant-level indices were then compared between HC and clinical participants using two-sample t-tests, with effect sizes reported as Cohen’s *d*.

#### Multivariate PLS Linking D-BART and CPT-3

To evaluate whether BART-derived metrics captured neurocognitive impulsivity as defined by the CPT-3, we conducted a Partial Least Squares (PLS) correlation analysis using a previously published implementation (myPLS toolbox https://github.com/MIPLabCH/myPLS). PLS is specifically designed to identify latent components (LCs) that maximize shared covariance between two sets of variables measured in the same participants.

The CPT-3 input matrix comprised the age-normed T-score variables described above (Detectability, Omissions, Commissions, Perseverations, Hit Reaction Time, Hit RT Standard Deviation, Variability, Hit RT Block Change, and Hit RT ISI Change). The D-BART input matrix combined traditional BART performance metrics (mean points, proportion of explosions, adjusted pumps, mean RT, RT standard deviation) with the four dynamic response-style parameters derived from mixed-effects modeling (RT-by-BI intercept, RT-by-BI slope, RT-by-DPC intercept, RT-by-DPC slope).

Prior to analysis, D-BART variables were corrected for age (including linear and quadratic terms) and sex using linear regression, in order to parallel the demographic normalization inherent in CPT-3 T-scores. Corrected variables were then mean-centered and scaled to unit variance to ensure comparable weighting across measures.

PLS was applied to the cross-covariance matrix between the CPT-3 and D-BART variable sets. For each latent component (LC), the procedure yields (i) a pair of variable-loading patterns—one for CPT-3 and one for D-BART—that define the multivariate signature of shared covariance, and (ii) a singular value quantifying the strength of this covariance. The proportion of total cross-block covariance explained by each LC was computed as the ratio of its singular value to the sum of singular values across all components.

The statistical significance of each LC was evaluated using non-parametric permutation testing (1,000 permutations). Specifically, the correspondence between CPT-3 and D-BART data matrices was randomly shuffled across participants, the PLS decomposition was recomputed for each permutation, and the observed singular value for each LC was compared against the resulting null distribution. An LC was considered significant if its observed covariance exceeded the 95th percentile of the permutation-based null distribution (p < .05).

The reliability of individual variable contributions (saliences) to each LC was assessed using bootstrap resampling (500 iterations). In each bootstrap sample, participants were resampled with replacement, the PLS model was recomputed, and the stability of each variable’s loading was estimated. Variables were considered to contribute reliably to a component if their bootstrap-derived confidence intervals did not cross zero.

Finally, participant-specific LC expression scores were computed by projecting each participant’s observed CPT-3 and D-BART data onto the corresponding LC loading vectors. These expression scores quantify the extent to which the multivariate behavioral pattern captured by a given LC is expressed in each individual and were used in subsequent diagnostic discrimination, correlation, and mediation analyses.

#### Evaluating the Diagnostic and Clinical Relevance of BART-Derived Response Patterns

Next, we evaluated whether CPT-3 and D-BART LC scores captured individual differences in vulnerability to real-world behavioral difficulties associated with neurocognitive impairment. Pearson correlation analyses were used to quantify associations between CRS-derived measures of behavioral difficulties and both CPT-3 and D-BART LC scores.

Given emerging evidence that behavioral manifestations of impulse control deficits may differ by sex, complementary analyses were conducted separately in male and female participants. In addition, we formally tested whether sex moderated the association between CPT-3 or D-BART LC scores and CRS outcomes using linear models including main effects of LC score and sex, as well as their interaction:

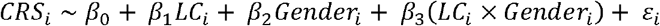

where *CRS*_*i*_denotes the outcome for individual *i*; *β*_*0*_is the intercept; *β*_*1*_the fixed effect of LC; *β*_*2*_the fixed effect of gender; *β*_*3*_the LC × gender interaction; and *ε*_*i*_ is the residual error term.

Moreover, mediation analyses were conducted to compare the relative ability of CPT-3 and D-BART scores to capture vulnerability to behavioral consequences associated with impulse control deficits. Specifically, we tested whether the association between CPT-3 LC scores and CRS behavioral measures was mediated by D-BART LC scores. Evidence for such mediation would support the added clinical value of the digital BART in capturing variance in real-world behavioral outcomes beyond that explained by traditional laboratory-based neurocognitive assessment.

Finally, to formally assess the contribution of repeated ecological BART assessments to the strength of cross-subject associations with clinical symptom measures, a systematic downsampling procedure was implemented. Analyses were restricted to the ADHD group, as only this group completed the 30-day ecological protocol with sufficient repeated assessments to permit controlled downsampling.

Firstly, assessment-level BART data were projected onto the previously derived D-BART-1 salience vector, yielding a latent D-BART-1 score for each ecological assessment. For each participant, a fixed proportion of available assessments (90%, 70%, 50%, 30%, or 10%) was randomly retained, and the remainder excluded. Random selection was repeated independently 1000 times for each availability threshold. At each repetition, participant-level D-BART-1 scores were recomputed as the mean of the retained assessment-level D-BART-

1 scores. Pearson correlations were then calculated across participants between the recomputed participant-level D-BART-1 scores and the Conners Hyperactivity/Impulsivity score.

A one-way ANOVA was used to test whether the mean correlation coefficient differed across availability thresholds. Post-hoc comparisons between adjacent thresholds were conducted using Welch-corrected two-sample *t*-tests, and effect sizes were quantified using Cohen’s *d*.

## Data Availability

All data produced in the present study are available upon reasonable request to the authors

## Notes

### Competing Interest Statement

The authors have declared no competing interest.

### Author Declarations

Commission Cantonale d'ethique de la recherche Geneve (CCER) gave ethical approval for this work. Basec ID: 2023-01061

### Summary of Updates

Figure 1 was revised to improve clarity and readability.

### Funding

- Swiss National Science Foundation: 212476
- Swiss National Science Foundation: 209096
- Swiss National Science Foundation: 162006
- Swiss National Science Foundation: 185897

## Bibliography

1. Moeller, F.G., et al., Psychiatric aspects of impulsivity. Am J Psychiatry, 2001. 158(11): p. 1783–93.

2. American Psychiatric, A., Diagnostic and Statistical Manual of Mental Disorders. DSM Library. 2013: American Psychiatric Association.-1.

3. Polanczyk, G., et al., The worldwide prevalence of ADHD: a systematic review and metaregression analysis. Am J Psychiatry, 2007. 164(6): p. 942–8.

4. Cortese, S., et al., Comparative efficacy and tolerability of medications for attention-deficit hyperactivity disorder in children, adolescents, and adults: a systematic review and network meta-analysis. Lancet Psychiatry, 2018. 5(9): p. 727–738.

5. Crisp, Z.C. and J.E. Grant, Impulsivity across psychiatric disorders in young adults. Compr Psychiatry, 2024. 130: p. 152449.

6. Perry, J.L. and M.E. Carroll, The role of impulsive behavior in drug abuse. Psychopharmacology (Berl), 2008. 200(1): p. 1–26.

7. Mungo, A., et al ., Impulsivity and its Therapeutic Management in Borderline Personality Disorder: a Systematic Review. Psychiatr Q, 2020. 91(4): p. 1333–1362.

8. Santana, R.P., et al., Impulsivity in Bipolar Disorder: State or Trait? Brain Sci, 2022. 12(10).

9. Lee, R.S.C., S. Hoppenbrouwers, and I. Franken, A Systematic Meta-Review of Impulsivity and Compulsivity in Addictive Behaviors. Neuropsychol Rev, 2019. 29(1): p. 14–26.

10. Robbins, T.W., et al., Neurocognitive endophenotypes of impulsivity and compulsivity: towards dimensional psychiatry. Trends Cogn Sci, 2012. 16(1): p. 81–91.

11. Zhong, G., et al., Transdiagnostic neuromodulation of impulsivity: current status and future trajectories. Transl Psychiatry, 2025. 15(1): p. 209.

12. Moffitt, T.E., et al., A gradient of childhood self-control predicts health, wealth, and public safety. Proc Natl Acad Sci U S A, 2011. 108(7): p. 2693–8.

13. Dalley, J.W., B.J. Everitt, and T.W. Robbins, Impulsivity, compulsivity, and top-down cognitive control. Neuron, 2011. 69(4): p. 680–94.

14. Karlsson Linner, R., et al., Multivariate analysis of1. 5 million people identifies genetic associations with traits related to self-regulation and addiction. Nat Neurosci, 2021. 24(10): p. 1367–1376.

15. Deng, W.Q., et al., Longitudinal characterization of impulsivity phenotypes boosts signal for genomic correlates and heritability. Mol Psychiatry, 2025. 30(2): p. 608–618.

16. Bezdjian, S., L.A. Baker, and C. Tuvblad, Genetic and environmental influences on impulsivity: a meta-analysis of twin, family and adoption studies. Clin Psychol Rev, 2011. 31(7): p. 1209–23.

17. He, X., et al., Impulsivity and non-suicidal self-injury in adolescents: a systematic review and meta-analysis of longitudinal studies. Front Psychiatry, 2025. 16: p. 1586922.

18. Read, R.W., et al., A study of impulsivity and adverse childhood experiences in a population health setting. Front Public Health, 2024. 12: p. 1447008.

19. Quinn, P. D. and K.P. Harden, Differential changes in impulsivity and sensation seeking and the escalation of substance use from adolescence to early adulthood. Dev Psychopathol, 2013. 25(1): p. 223–39.

20. French, B., et al., The impacts associated with having ADHD: an umbrella review. Front Psychiatry, 2024. 15: p. 1343314.

21. Biederman, J., Attention-deficit/hyperactivity disorder: a selective overview. Biol Psychiatry, 2005. 57(11): p. 1215–20.

22. Shaw, M., et al., A systematic review and analysis of long-term outcomes in attention deficit hyperactivity disorder: effects of treatment and non-treatment. BMC Med, 2012. 10: p. 99.

23. Hofmann, W., et al., Self-control and behavioral public policy. Curr Opin Psychol, 2024. 59: p. 101873.

24. Duckworth, A.L., K.L. Milkman, and D. Laibson, Beyond Willpower: Strategies for Reducing Failures of Self-Control. Psychol Sci Public Interest, 2018. 19(3): p. 102–129.

25. Dalley, J.W. and T.W. Robbins, Fractionating impulsivity: neuropsychiatric implications. Nat Rev Neurosci, 2017. 18(3): p. 158–171.

26. Beck, L.H., et al., A continuous performance test of brain damage. J Consult Psychol, 1956. 20(5): p. 343–50.

27. Albrecht, B., et al ., ADHD History of the Concept: the Case of the Continuous Performance Test. Current Developmental Disorders Reports, 2015. 2(1): p. 10–22.

28. Nguyen, R., et al., Behavioral measures of state impulsivity and their psychometric properties: A systematic review. Personality and Individual Differences, 2018. 135: p. 67–79.

29. Esteves, M., et al., Assessing Impulsivity in Humans and Rodents: Taking the Translational Road. Front Behav Neurosci, 2021. 15: p. 647922.

30. Bechara, A., et al., Insensitivity to future consequences following damage to human prefrontal cortex. Cognition, 1994. 50(1-3): p. 7–15.

31. Lejuez, C.W., et al., Evaluation of a behavioral measure of risk taking: the Balloon Analogue Risk Task (BART). J Exp Psychol Appl, 2002. 8(2): p. 75–84.

32. Green, L. and J. Myerson, How many impulsivities? A discounting perspective. J Exp Anal Behav, 2013. 99(1): p. 3–13.

33. MacLean, R.R., et al., Extending the Balloon Analogue Risk Task to Assess Naturalistic Risk Taking via a Mobile Platform. J Psychopathol Behav Assess, 2018. 40(1): p. 107–116.

34. Bari, A. and T.W. Robbins, Inhibition and impulsivity: behavioral and neural basis of response control. Prog Neurobiol, 2013. 108: p. 44–79.

35. Peterson, B.S., et al., Tools for the Diagnosis of ADHD in Children and Adolescents: A Systematic Review. Pediatrics, 2024. 153(4).

36. Buelow, M.T., B.M. Okdie, and J.M. Kowalsky, Ecological validity of common behavioral decision making tasks: evidence across two samples. J Clin Exp Neuropsychol, 2024. 46(3): p. 187–206.

37. Chaytor, N. and M. Schmitter-Edgecombe, The Ecological Validity of Neuropsychological Tests: A Review of the Literature on Everyday Cognitive Skills. Neuropsychology Review, 2003. 13(4): p. 181–197.

38. Carr, T.H., C.M. Arrington, and S.M. Fitzpatrick, Integrating cognition in the laboratory with cognition in the real world: the time cognition takes, task fidelity, and finding tasks when they are mixed together. Front Psychol, 2023. 14: p. 1137698.

39. Beauchaine, T.P., A.R. Zisner, and C.L. Sauder, Trait Impulsivity and the Externalizing Spectrum. Annu Rev Clin Psychol, 2017. 13: p. 343–368.

40. Guarana, C.L., et al., Sleep and self-control: A systematic review and meta-analysis. Sleep Med Rev, 2021. 59: p. 101514.

41. Logan, R.W., et al., Impact of Sleep and Circadian Rhythms on Addiction Vulnerability in Adolescents. Biol Psychiatry, 2018. 83(12): p. 987–996.

42. Bandarabadi, M., et al., Inactivation of hypocretin receptor-2 signaling in dopaminergic neurons induces hyperarousal and enhanced cognition but impaired inhibitory control. Mol Psychiatry, 2024. 29(2): p. 327–341.

43. Pearlstein, J.G., et al., Neurocognitive mechanisms of emotion-related impulsivity: The role of arousal. Psychophysiology, 2019. 56(2): p. e13293.

44. Thunberg, C., et al., On the (un)reliability of common behavioral and electrophysiological measures from the stop signal task: Measures of inhibition lack stability over time. Cortex, 2024. 175: p. 81-105.

45. Roos, C.R. and K. Witkiewitz, A contextual model of self-regulation change mechanisms among individuals with addictive disorders. Cl in Psychol Rev, 2017. 57: p. 117–128.

46. Sobolev, M., et al., The Digital Marshmallow Test (DMT) Diagnostic and Monitoring Mobile Health App for Impulsive Behavior: Development and Validation Study. JMIR Mhealth Uhealth, 2021. 9(1): p. e25018.

47. Moore, R.C., J. Swendsen, and C.A. Depp, Applications for self-administered mobile cognitive assessments in clinical research: A systematic review. Int J Methods Psychiatr Res, 2017. 26(4).

48. Schneider, M., et al., Psychiatric disorders from childhood to adulthood in 22q11. 2 deletion syndrome: results from the International Consortium on Brain and Behavior in 22q11. 2 Deletion Syndrome. Am J Psychiatry, 2014. 171(6): p. 627–39.

49. Logan, G.D., et al., On the ability to inhibit thought and action: general and special theories of an act of control. Psychol Rev, 2014. 121(1): p. 66–95.

50. Schall, J.D., T.J. Palmeri, and G.D. Logan, Models of inhibitory control. Philos Trans R Soc Lond B Biol Sci, 2017. 372(1718).

51. Diederich, A. and J.S. Trueblood, A dynamic dual process model of risky decision making. Psychol Rev, 2018. 125(2): p. 270–292.

52. Pleskac, T.J. and A. Wershbale, Making assessments while taking repeated risks: a pattern of multiple response pathways. J Exp Psychol Gen, 2014. 143(1): p. 142–62.

53. Krajbich, I., et al., Rethinking fast and slow based on a critique of reaction-time reverse inference. Nat Commun, 2015. 6: p. 7455.

54. Kahneman, D.a., Thinking, fast and slow. 2011: 1st ed. New York : Farrar, Straus and Giroux, [2011] ©2011.

55. Coon, J. and M.D. Lee, A Bayesian method for measuring risk propensity in the Balloon Analogue Risk Task. Behav Res Methods, 2022. 54(2): p. 1010–1026.

56. Zhou, R. and M.A. Pitt, Dual-process modeling of sequential decision making in the balloon analogue risk task. Cogn Psychol, 2024. 149: p. 101629.

57. Zhou, R., J.I. Myung, and M.A. Pitt, The scaled target learning model: Revisiting learning in the balloon analogue risk task. Cogn Psychol, 2021. 128: p. 101407.

58. Egeland, J. and I. Kovalik-Gran, Measuring several aspects of attention in one test: the factor structure of conners’s continuous performance test. J Atten Disord, 2010. 13(4): p. 339–46.

59. Hartman, C.A., et al., Anxiety, mood, and substance use disorders in adult men and women with and without attention-deficit/hyperactivity disorder: A substantive and methodological overview. Neurosci Biobehav Rev, 2023. 151: p. 105209.

60. Roncero, C., et al., Gender differences in ADHD and impulsivity among alcohol or alcohol- and cocaine-dependent patients. Front Psychiatry, 2025. 16: p. 1446970.

61. Cortese, S., et al., Gender differences in adult attention-deficit/hyperactivity disorder: results from the National Epidemiologic Survey on Alcohol and Related Conditions (NESARC). J Clin Psychiatry, 2016. 77(4): p. e421–8.

62. Faheem, M., et al., Gender-based differences in prevalence and effects of ADHD in adults: A systematic review. Asian J Psychiatr, 2022. 75: p. 103205.

63. Kok, F.M., et al., Problematic Peer Functioning in Girls with ADHD: A Systematic Literature Review. PLoS One, 2016. 11(11): p. e0165119.

64. Case, A. and A. Deaton, Mortality and morbidity in the 21(st) century. Brookings Pap Econ Act, 2017. 2017: p. 397–476.

65. Richmond-Rakerd, L.S., et al., Clustering of health, crime and social-welfare inequality in 4 million citizens from two nations. Nat Hum Behav, 2020. 4(3): p. 255–264.

66. Chaytor, N. and M. Schmitter-Edgecombe, The ecological validity of neuropsychological tests: a review of the literature on everyday cognitive skills. Neuropsychol Rev, 2003. 13(4): p. 181–97.

67. Wallsten, T.S., T.J. Pleskac, and C.W. Lejuez, Modeling behavior in a clinically diagnostic sequential risk-taking task. Psychol Rev, 2005. 112(4): p. 862–80.

68. Wang, J., et al., Genome-wide association studies (GWAS) and post-GWAS analyses of impulsivity: A systematic review. Prog Neuropsychopharmacol Biol Psychiatry, 2024. 132: p. 110986.

69. Courtney, K.E., et al., The relationship between measures of impulsivity and alcohol misuse: an integrative structural equation modeling approach. Alcohol Clin Exp Res, 2012. 36(6): p. 923–31.

70. Mazumder, A., S. Erb, and M.A. Fournier, State impulsivity and substance use: A systematic review and meta-analysis protocol. PLoS One, 2026. 21(4): p. e0346779.

71. Siddi, F., et al., Mobile health and neurocognitive domains evaluation through smartphones: A meta-analysis. Comput Methods Programs Biomed, 2021. 212: p. 106484.

72. Pinto, J. O., et al., Ecological validity in neurocognitive assessment: Systematized review, content analysis, and proposal of an instrument. Appl Neuropsychol Adult, 2025. 32(2): p. 577–594.

73. Jonas, R.K., C.A. Montojo, and C.E. Bearden, The 22q11. 2 deletion syndrome as a window into complex neuropsychiatric disorders over the lifespan. Biol Psychiatry, 2014. 75(5): p. 351–60.

74. Sussman, C.J., et al., Internet and Video Game Addictions: Diagnosis, Epidemiology, and Neurobiology. Child Adolesc Psychiatr Clin N Am, 2018. 27(2): p. 307–326.

75. Tran, L.T., et al., The prevalence of gambling and problematic gambling: a systematic review and meta-analysis. Lancet Public Health, 2024. 9(8): p. e594–e613.

76. Qi, J., Y. Yan, and H. Yin, Screen time among school-aged children of aged 6-14: a systematic review. Glob Health Res Policy, 2023. 8(1): p. 12.

77. Sohn, S.Y., et al., Prevalence of problematic smartphone usage and associated mental health outcomes amongst children and young people: a systematic review, meta-analysis and GRADE of the evidence. BMC Psychiatry, 2019. 19(1): p. 356.

78. Meng, S.Q., et al., Global prevalence of digital addiction in general population: A systematic review and meta-analysis. Clin Psychol Rev, 2022. 92: p. 102128.

79. Shiferaw, B.D., et al., Impact of digital addiction on youth health: A systematic review and meta-analysis. J Behav Addict, 2025. 14(3): p. 1129–1158.

80. Becker, T.D. and T.R. Rice, Youth vaping: a review and update on global epidemiology, physical and behavioral health risks, and clinical considerations. Eur J Pediatr, 2022. 181(2): p. 453–462.

81. Kim, M.S., et al., Association of genetic risk, lifestyle, and their interaction with obesity and obesity-related morbidities. Cell Metab, 2024. 36(7): p. 1494–1503 e3.

82. Greenberg, N.R., et al., Difficulties in impulse control in adolescents with problematic use of the internet and self-injurious behaviors. Psychiatry Res, 2022. 317: p. 114919.

83. Intravia, J., A.G. Vito, and G.C. Rocheleau, Low Self-Control and Vaping: The Mediating Role of Health and Risk Perceptions. Subst Use Misuse, 2022. 57(6): p. 956–966.

84. Mao, B., et al., Future Time Perspective and Bedtime Procrastination: The Mediating Role of Dual-Mode Self-Control and Problematic Smartphone Use. Int J Environ Res Public Health, 2022. 19(16).

85. Cudo, A., et al., Dysfunction of Self-Control in Facebook Addiction: Impulsivity Is the Key. Psychiatr Q, 2020. 91(1): p. 91–101.

86. Liu, Y., et al., Self-control as a mediator of age on students’ compulsive buying. Psych J, 2022. 11(2): p. 259–262.

87. Sandini, C., et al., Development of Structural Covariance From Childhood to Adolescence: A Longitudinal Study in 22q11. 2DS. Front Neurosci, 2018. 12: p. 327.

88. Schaer, M., et al., Deviant trajectories of cortical maturation in 22q11. 2 deletion syndrome (22q11DS): a cross-sectional and longitudinal study. Schizophr Res, 2009. 115(2-3): p. 182–90.

89. American Psychiatric Association, Diagnostic and Statistical Manual of Mental Disorders. 5th ed. 2013, Washington, DC.

90. Kaufman, J., et al., Schedule for Affective Disorders and Schizophrenia for School-Age Children-Present and Lifetime Version (K-SADS-PL): initial reliability and validity data. J Am Acad Child Adolesc Psychiatry, 1997. 36(7): p. 980–8.

91. Conners, C.K., J. Pitkanen, and S.R. Rzepa, Conners 3rd Edition (Conners 3; Conners 2008), in Encyclopedia of Clinical Neuropsychology, J.S. Kreutzer, J. DeLuca, and B. Caplan, Editors. 2011, Springer New York: New York, NY. p. 675–678.

92. Keith Conners, C., G. Sitarenios, and L.E. Ayearst, Conners’ Continuous Performance Test Third Edition, in Encyclopedia of Clinical Neuropsychology, J.S. Kreutzer, J. DeLuca, and B. Caplan, Editors. 2018, Springer International Publishing: Cham. p. 929–933.

93. Riccio, C.A., et al., Validity of the Auditory Continuous Performance Test in differentiating central processing auditory disorders with and without ADHD. J Learn Disabil, 1996. 29(5): p. 561–6.

94. Culbertson, W.C. and E.A. Zillmer, The Tower of London(DX): a standardized approach to assessing executive functioning in children. Arch Clin Neuropsychol, 1998. 13(3): p. 285–301.

95. D’Elia, L.F.S., P., Uchiyama, C.L., & White, T., Color Trails Test: Professional Manual. Odessa, FL: Psychological Assessment Resources., 1989.

96. Williams, J., et al., Children’s color trails. Arch Clin Neuropsychol, 1995. 10(3): p. 211–23.

97. d, W., The Wechsler intelligence scale for children—third edition: administration and scoring manual. 1991, San Antonio: Psychological corporation.

98. D., W., Wechsler adult intelligence scale-III: administration and scoring manual. 1997, San Antonio: Psychological Corporation.

99. Torous, J. and A. Vaidyam, Multiple uses of app instead of using multiple apps - a case for rethinking the digital health technology toolbox. Epidemiol Psychiatr Sci, 2020. 29: p. e100.

