## Supplementary_Material for "Reconciling neurocognitive and behavioral impulsivities through ecological assessment and multivariate modelling of cognitive control dynamics"

### Supplementary Analysis 1

#### Methods

In the main text, group differences between healthy controls (HCs) and clinical participants (CPs) were identified using Dynamic RT parameters derived from within-task response-time dynamics. Specifically, these indices quantified how RT changed as a function of Balloon Inflation Number, which indexed objective risk, and Distance to Point Collection, which indexed subjective uncertainty. Compared with CPs, HCs showed greater RT slowing as objective risk increased, longer RTs immediately prior to cash-out, and sharper RT acceleration as uncertainty decreased. The aim of the present supplementary analysis was to determine whether similarly group differences could also be detected using conventional participant-level BART summary measures. For each participant, we therefore computed explosion rate, RT standard deviation, adjusted mean points, mean total points, and mean RT. Group differences between HCs and CPs were then assessed using two-sample *t*-tests.

#### Figure 1

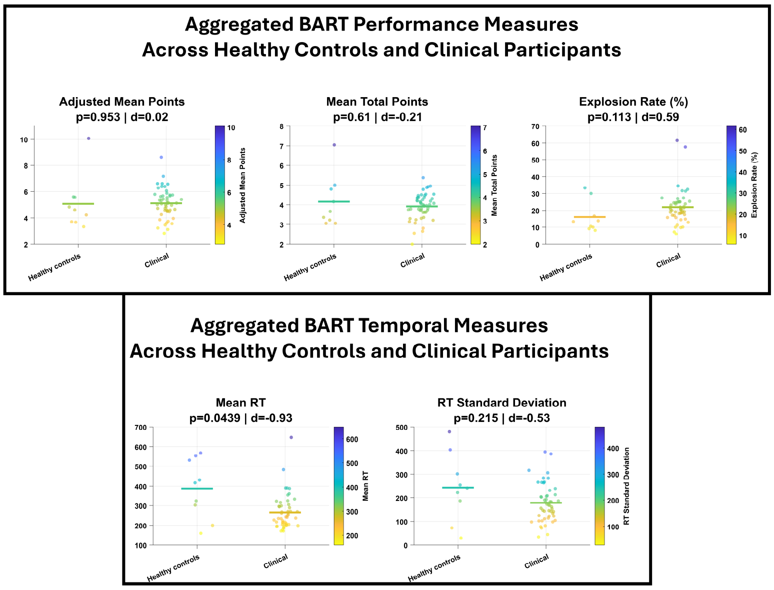


Traditional aggregated BART measures in Healthy Controls and the pooled Clinical group. Top panels: Distribution of participant-level aggregated BART performance measures, including adjusted mean points, mean total points, and explosion rate. Bottom panels: Distribution of participant-level aggregated BART temporal measures, including mean RT and RT standard deviation. Scatterplots show the distribution of individual values in Healthy Controls and the pooled Clinical group, with each point representing one participant. Color coding reflects the value of the corresponding measure in each panel. The horizontal bar indicates the group mean and is colored according to its corresponding value. Panel titles report the p-value of the two-sample t-test and Cohen’s d for the group comparison.

#### Results

Traditional BART summary measures showed limited sensitivity to group differences. Healthy controls (HCs) and clinical participants (CPs) did not differ significantly in explosion rate ($p$ = .113, $d$ = 0.59), RT standard deviation ($p$ = .215, $d$ = -0.53), adjusted mean points ($p$ = .953, $d$ = 0.02), or mean total points ($p$ = .610, $d$ = -0.21) (Supplementary Figure S1). A significant group difference was observed only for mean RT, which was higher in HCs than in CPs ($p$ = .0439, $d$ = -0.93), consistent with the main-text result showing that reduced RT modulation in CPs contributed to a smaller overall increase in mean RT relative to HCs. Together, these results illustrate the added value of Dynamic RT parameters, which captured group differences that were largely not detectable using conventional participant-level BART summary measures.
